# Prevalence and determinants of peripheral arterial disease in children with nephrotic syndrome

**DOI:** 10.1101/2022.03.21.22272734

**Authors:** Gbenga Akinyosoye, Adaobi U Solarin, Adeyemi Dada, Motunrayo O Adekunle, Alaba B Oladimeji, Adeola O Owolabi, Clement M Akinsola, Barakat A Animasahun, Fidelis O Njokanma

## Abstract

Peripheral arterial disease (PAD) is the least studied complication of nephrotic syndrome (NS). Risk factors which predispose children with NS to developing PAD include hyperlipidaemia, hypertension and prolonged use of steroids. The development of PAD significantly increases the morbidity and mortality associated with NS as such children are prone to sudden cardiac death. The ankle brachial index (ABI) is a tool that has been proven to have high specificity and sensitivity in detecting PAD even in asymptomatic individuals. We aimed to determine the prevalence of PAD in children with NS and to identify risk factors that can independently predict its development. A comparative cross-sectional study was conducted involving 200 subjects (100 with NS and 100 apparently healthy comparative subjects that were matched for age, sex and socioeconomic class). Systolic blood pressures were measured in all limbs using the pocket Doppler machine (Norton Doppler scan machine). ABI was calculated as a ratio of ankle to arm systolic blood pressure. PAD was defined as ABI less than 0.9. The prevalence of PAD was significantly higher in children with NS than matched comparison group (44.0% vs 6.0%, p < 0.001). Average values of waist and hip circumference were significantly higher in subjects with PAD than those without PAD (61.68± 9.1cm and 67.6± 11.2 cm vs 57.03 ± 8.3cm and 65.60± 12.5cm respectively, p< 0.005). Serum lipids (triglyceride, very low density lipoprotein, total cholesterol and low density lipoprotein) were also significantly higher in subjects with PAD than those without PAD [106.65mg/dl (67.8-136.7) vs 45.72mg/dl (37.7-61.3), 21.33mg/dl (13.6-27.3) vs 9.14mg/dl (7.5-12.3), 164.43mg/dl (136.1-259.6) vs 120.72mg/dl (111.1-142.1) and 93.29mg/dl (63.5-157.3) vs 61.84mg/dl (32.6-83.1), respectively p< 0.05]. Increasing duration since diagnosis of NS, having a steroid resistant NS and increasing cumulative steroid dose were independent predictors of PAD in children with NS; p< 0.05 respectively).

## Introduction

Nephrotic syndrome (NS) remains the most common manifestation of glomerular disease in childhood.[1-3] It is defined by massive proteinuria, hypoalbuminaemia, generalized oedema and hyperlipidaemia.[4] Most studies estimate the global incidence of NS to be 2 to 7 per 100,000 population with a prevalence of 16 per 100,000 population.[5] Children with NS are at increased risk of complications such as infections, acute kidney injury, thromboembolism and peripheral arterial disease (PAD).[5,6] However, of these complications, PAD remains the least studied, especially in children. Peripheral arterial disease is an umbrella term used to describe a group of disorders that result in structural and functional alteration of arteries supplying blood to the viscera and limbs. It is a circulatory problem in which narrowed arteries from widespread accumulation of fatty deposits reduce blood flow to the heart, brain, and legs.[7] Peripheral arterial disease has been established to be prevalent among children with chronic kidney disease, especially children with NS, with a prevalence as high as 80% documented in Egypt by Mohammed et al.[4] The development of PAD can become progressive and associated with a significant increase in cardiovascular related events like cardiac arrest, arrhythmias, transient ischaemic attack and sudden cardiac death that can occur in children.[8,9]

There is a paucity of data on the burden of PAD in children with NS, with the few studies reporting a high prevalence of PAD, above 80%.[4,8] Thus, with a prevalence of PAD in children with NS as high as 80% compared to 9% in apparently healthy controls in an African study done in Egypt by Mohammed et al[4] the need for prompt screening of children with NS for PAD cannot be overemphasized. Furthermore, children with NS who develop PAD generally have a worse outcome and survival than those without PAD.[10]

Nephrotic syndrome is associated with hyperlipidaemia which predisposes to PAD through endothelial dysfunction as well as inflammation.[8,11] High levels of lipid in the blood promote plaque formation which can accumulate in the arteries and cause narrowing of the lumen of these arteries and thus limit blood flow. Other factors that contribute to the development of PAD in children with NS include hypertension, prolonged use of steroids, uncontrolled hyperglycaemia and proteinuria.[4,12] These risk factors are associated with NS and its treatment, and can lead to the development of PAD by causing endothelial damage. Children with NS and associated PAD are significantly at an increased risk of thromboembolic phenomena such as cerebrovascular accidents, compared to other children with NS who do not have PAD and to the general population.[13]

Nephrotic syndrome coexisting with PAD is also a known risk factor for myocardial infarction in adulthood, hence children with the co-morbidity are likely to develop adverse cardiovascular events at a relatively young age compared to their healthy counterparts.[14,15] One consistent risk factor from previous studies for the development of PAD is atherosclerosis.[9,16-20] Hyperlipidaemia is a known risk factor for the development of atherosclerosis, and is a major component of NS that predisposes to vascular damage and, as a result, poses a risk for the development of PAD, particularly in children with NS.[8,11] A valid and reliable screening tool in detecting PAD is the ankle brachial index (ABI). The ABI is proven to be a sensitive, non-invasive, user friendly, readily available, and cost-effective screening tool for PAD that can be used in children.[4,21] The ABI is defined as the ratio of systolic blood pressure measured at the ankle to that measured at the brachial artery.[22] Normal values for ABI are between 0.9 and 1.3.[21] An abnormal value below 0.9 is a powerful independent marker of cardiovascular risk, especially atherosclerosis.[20,23] A three-to-four-fold increase in cardiac associated mortalities arises when the ABI value is less than 0.9.[23] Apart from its ability to detect the presence of PAD, it also helps in prognosticating cardiovascular events and functional impairments even in the absence of symptoms.[23-25] Studies have consistently shown the ABI to have a minimum sensitivity and specificity of 90% and 98% respectively in predicting PAD, and with a minimum positive predictive value and negative predictive value of 90% and 98.63% respectively.[22-29]

Despite the high burden of childhood NS, data on prevalence of PAD in children with NS is scarce. The study done in Egypt by Mohammed et al[4] among a cohort of children with NS recruited only those with steroid resistant NS, whereas the current study recruited all children with NS, irrespective of their steroid status. Environmental factors and the effect of genetic composition may account for variations in steroid response among children with NS in Egypt as compared to other parts of the continent.[4] We aimed to determine the burden of PAD in children with NS and identify factors that can independently predict the development of PAD. With this being achieved, it is recommended that screening for PAD in children with NS will be useful to prevent cardiovascular complications before they arise.

## Materials and Methods

The study described in this report was carried out at the paediatric nephrology clinic of Lagos State University Teaching Hospital (LASUTH) Ikeja, a tertiary centre that receives referrals from all parts of Lagos state and environs. Lagos is a metropolitan state with a heterogeneous population, situated in the coastal region of South-Western Nigeria. It has a population of about 9 million people, according to the last national population census carried out in 2006, and it was projected to have increased to over 12 million by 2016.[30] Though Lagos is cosmopolitan, the majority of its inhabitants are Yorubas. The paediatric nephrology clinic of LASUTH runs on a weekly basis and attends to diverse cases. On average, about eight patients with nephrotic syndrome are seen per week, most of them being follow up cases. However, some patients with acute complaints are admitted to the ward via the paediatric emergency unit.

### Study Design

This was a comparative cross-sectional study involving children with NS as well as age, sex and socioeconomic class matched apparently healthy participants in the comparison group. A group matching was done after recruitment of the cases.

### Study Population

This comprised 100 patients with NS aged one to fifteen years attending the Paediatric Nephrology Clinic of LASUTH. Hundred apparently healthy children with no history of renal, haematological and cardiovascular diseases were matched for age, sex and social class to serve as comparison group. The participants in the comparative group were recruited from the paediatric out-patient department, the immunization clinic of LASUTH, children attending specialist clinics such as the dermatology clinic for follow up and teenage children presenting for medical fitness for school resumption.

### Inclusion Criteria

All children with a diagnosis of NS aged 1 to 15 years were recruited as the cases and the comparative group had children aged 1 to 15 years with no fever or acute illness.

### Exclusion Criteria

For the cases, children with diabetes mellitus, sickle cell anaemia, underlying cardiac abnormalities and previous renal abnormalities other than NS were excluded. In the comparison group, in addition to these, chidren with fever, asthma, proteinuria greater than 1+, obese children and those on steroids were excluded.

### Data Collection Procedures

The researchers obtained data from all participants using a self pre-designed proforma. The proforma included sections for the participants bio-data (age, sex and ethnic group), date of diagnosis of NS, presence of any other chronic illness e.g., cardiac or, renal disease, sickle cell anaemia, etc. Social history of passive smoking, socio economic status and family history of cardiac and renal diseases were obtained. Use and duration of medications especially steroids and anti-hypertensives were documented. The medical records of participants were also checked to ascertain the cumulative dose of steroid used and identify those who have had a renal biopsy. Their response to steroid was documented from the case note into steroid sensitive (remission achieved with steroid therapy alone),[31] steroid dependent (two consecutive relapses occurring during corticosteroids tapering or within fourteen days of its cessation)[31,32] and steroid resistant (failure to achieve response in spite of eight weeks of oral prednisolone at 60mg/m^2^/day).[31,32] Each participant had a general physical examination done to rule out any asymptomatic abnormality for example, a cardiac murmur.

**ANKLE BRACHIAL INDEX** was measured using appropriate blood pressure cuffs (bladder width at least 40% of the limb circumference, length encircling 80% to 100% of the limb circumference) and pocket doppler probe electronics.[24,33] Subjects were positioned supine for 5 minutes.[24] The systolic blood pressure at the brachial artery of each arm, the posterior tibial artery and dorsalis pedis artery sof each ankle were measured. The higher of the two upper limb pressures and of the lower limb pressures were used to calculate the ABI. The ratio of the systolic ankle pressure to the systolic brachial pressure gives the ABI.[33] ABI was classified as low (< 0.9), normal (0.9 - 1.3), and high (> 1.3).[4,34,35] The following steps were taken by the investigators in performing the ABI measurement using the pocket doppler.

1. Following explanation of the procedure, the participant was made to lie supine for five minutes in a quiet room, removing socks, shoes and tight clothing.
2. The pocket doppler was assembled and the investigator donned on clean gloves.
3. An appropriate sized blood pressure cuff was tied on the upper arm approximately one to two centimetres above the ante-cubital fossa.
4. The brachial pulse was identified and gel was applied over the pulsation.
5. Doppler was turned on and the probe was held at 45 degree angle towards the blood flow.
6. The probe was moved slowly through the gel in circular motion until a pulse sound was heard.
7. The blood pressure cuff was inflated until the pulse sound disappeared. It was then further inflated by 10-20mmHg.
8. The cuff was gradually deflated until the arterial sound returned. The pressure at which the sound returned was noted and the cuff completely deflated.
9. Steps 3-7 were repeated on the other arm. The higher reading of the two systolic arm pressures was used to calculate the ABI.
10. The dorsalis pedis and posterior tibial pulsations were identified. With a similar method outlined in steps 3-7, the doppler machine was used to determine the systolic blood pressures. The highest reading of the four systolic pulses was used to calculate the ABI.
11. Gel was cleaned off the subject.
12. ABI was calculated as the highest of the four systolic pressures from the two ankles divided by the higher of the two brachial pressures and the value documented.
13. The participant was thanked and assisted to dress up.

#### BLOOD AND URINE SAMPLES COLLECTION

Five millilitres (5mls) of blood for serum protein, albumin and lipid profile was collected in a lithium heparin bottle. A drop of blood was put on the blood glucose meter to check for the random blood glucose which was subsequently recorded. Freshly voided urine was collected in a universal bottle and urinalysis was done using Siemens urinalysis strips Combi-9® with the remaining urine sample analysed for protein and creatinine to determine a spot protein-creatinine ratio.

#### BIOCHEMICAL ANALYSIS

Blood and urine samples were collected by the researchers and transported immediately to the research laboratory. The blood samples were immediately centrifuged by the researcher and stored. Analysis of the samples were done by the chemical pathologist and assisted by the researchers. Blood samples were centrifuged at 350 revolutions per minute (rpm) for 5 minutes; and the serum separated and stored at a temperature of (−20^0^C) in a refrigerator until each batch of samples were collected and afterwards analysed. Lagos State University Teaching Hospital (LASUTH) enjoys un-interrupted power supply largely from the independent power project (IPP) situated in Alausa, Ikeja and therefore maintenance of sample at required temperature expected was achieved.

Serum cholesterol was analysed using enzymatic (Cholesterol oxidase) colorimetric method.[36] Serum triglyceride was analysed using enzymatic (glycerol kinase) colorimetric method.[37] Total serum protein was analysed using the Biuret method. Serum albumin was analysed using colorimetric method (Bromocresol Green).

Siemens (Combi-9®) urinary test strips was used for urinalysis assessment. Urine protein was analysed using colorimetric method (Pyrogallol Red).

### Data Analysis

Data was analysed using Microsoft Excel and Statistical Package for Social Sciences (SPSS) version 24.0 (IBM, Inc, Chicago, Illinios USA). Demography of participants were presented as frequency and percentages. Tables and figures were used to present the variables as appropriate.

Bar chart was used to present prevalence of peripheral arterial disease (which is the dependent variable/outcome) among patients with NS and the comparison group. Fisher’s exact test was used to compare prevalence of PAD between the two groups. Association between categorical independent variables (sex, socio-economic class, age group, BMI for age Z score categories) and outcome variable (PAD) was assessed using Pearson’s chi square or Fisher’s exact test for expected frequencies less than five.

For numeric independent variables (e.g. weight, blood pressure, lipid profile, random blood glucose), Kolmogorov-Smirnov test was used to assess data normality. Independent student t-test was used to compare means according to the presence or absence of PAD when normally distributed. Mann Whitney U test was used for comparison of the median values of two groups when data were skewed (not normally distributed).

Multiple logistic regression, a form of multi-variate analysis was used to determine independent predictors of PAD among NS participants among a list of covariates (socio-demographics, clinical and biochemical parameters). Probability (p) value was considered significant at less than 0.05 and at a confidence interval of 95%.

### Ethical Approval

Approval was granted by the Health and Research Ethics committee of LASUTH with approval number LREC/06/10/1173. The personal details of the patients were used in a non-identifiable and confidential manner. Written informed consent was obtained from the caregivers and in addition assent from children seven years and above.

## Results

### Demographic, clinical and laboratory characteristics

One hundred (100) children aged 1-15 years with NS and 100 age, sex and social class matched comparison group were recruited for the study. The mean age for participants with nephrotic syndrome and the comparison group were 7.53 ± 2.6 years and 7.07 ± 2.6 years, respectively. The predominant age group was 6 to 10 years. Hip circumference, WAZ, HAZ, BAZ were comparable between the two groups (p>0.05). Waist circumference, waist hip ratio and lipid profile indices were signifcantly higher among children with NS than the comparison group. Four fifth of all the NS children were steroid sensitive. Average duration since diagnosis of NS was 2 years with a median cummulative steroid dose of about 4000mg as shown in table 1.

**Table 1:** Socio-demographic, clinical and laboratory characteristics of participants.

| Variables | Case<br>(n = 100)<br>n (%) | Comparative<br>group (n = 100)<br>n (%) | Total | $\chi^2$ | p-value |
| --- | --- | --- | --- | --- | --- |
| **Age group (Years)** |  |  |  |  |  |
| 1 - 5 | 26(26.0) | 26(26.0) | 52(26.0) | 0.000 | 1.000 |
| 6 - 10 | 44(44.0) | 44(44.0) | 88(44.0) |  |  |
| 11 - 15 | 30(30.0) | 30(30.0) | 60(30.0) |  |  |
| Mean ± SD | 7.53±2.6 | 7.07±2.6 | 7.30±2.6** | 0.902 | 0.368 |
| **Gender** |  |  |  |  |  |
| Male | 58(58.0) | 58(58.0) | 116(58.0) | 0.000 | 1.000 |
| Female | 42(42.0) | 42(42.0) | 84(42.0) |  |  |
| **Social class** |  |  |  |  |  |
| Lower | 41(41.0) | 41(41.0) | 82(41.0) | 0.000 | 1.000 |
| Middle | 36(36.0) | 36(36.0) | 72(36.0) |  |  |
| Upper | 23(23.0) | 23(23.0) | 46(23.0) |  |  |
| Waist circumference | 61.68±9.1 | 57.03±8.3 | 59.36±9.0** | 0.902 | <0.001* |
| Hip circumference | 67.65±11.2 | 65.60±12.5 | 66.63±11.9** | 3.766 | 0.224 |
| Waist hip ratio | 0.37±1.3 | 0.32±1.1 | 0.89±0.1** | 1.221 | <0.001* |
| WAZ | 0.37±1.3 | 0.32±1.1 | 0.35±1.2** | 3.842 | 0.831 |
| HAZ | -1.06±1.3 | -0.77±1.2 | -0.91±1.3** | 1.373 | 0.172 |
| BAZ | 1.27±1.3 | 1.02±1.1 | 1.15±1.2** | 1.204 | 0.231 |
| **RBG (mg/dl)** | 85.71±17.1 | 86.87±19.5 | 86.29±18.3** | -0.447 | 0.656 |
| **Serum protein(mg/dl)** | 5.30±1.0 | 7.09±1.0 | 6.20±1.3** | 12.706 | <0.001* |
| **Albumin (mg/dl)** | 3.45±0.9 | 4.63±0.5 | 4.04±0.9** | 11.242 | <0.001* |
| **Triglyceride (mg/dl)** | 106.7 (67-137) | 45.7 (38-61) | 63.7(42-115) | -5.122 | <0.001* |
| **VLDL (mg/dl)** | 21.3(14-27) | 9.1(8-12) | 12.5(8-22) | -4.848 | <0.001* |
| **TC (mg/dl)** | 164.4(136-259) | 120.7(111-142) | 139(113-169) | -4.534 | <0.001* |
| **LDL (mg/dl)** | 93.3(63-157) | 61.8(32-83) | 72.6(44-111) | -4.242 | <0.001* |
| **Duration since diagnosis (Months)** | 24.0(12.51) | NA | NA |  |  |
| **Cumulative steroid dose (mg)** | 4165(3162-5280) | NA | NA |  |  |
| **Duration of steroid use (months)** | 12.0 (10.0-36.0) | NA | NA |  |  |
| **Response to steroid** |  |  |  |  |  |
| Steroid dependent | 3(3.0) | NA | NA |  |  |
| Steroid resistance | 17(17.0) | NA | NA |  |  |
| Steroid sensitive | 80(80.0) | NA | NA |  |  |

**Table 2:** Mean ABI in cases and comparison group.

| **Variables** | **Case (n = 100)**<br>Mean ± SD | **Comparison group (n = 100)**<br>Mean ± SD | **t-value** | **p-value** |
| --- | --- | --- | --- | --- |
| **Systolic ankle pressure** |  |  |  |  |
| Mean ± SD | 90.7 ± 24.4 | 102.7 ± 24.4 | 6.421 | < 0.001* |
| Range | 70.0 – 130.0 | 70.0 – 130.0 |  |  |
| **Systolic brachial pressure** |  |  |  |  |
| Mean ± SD | 102.2 ± 26.3 | 98.7 ± 22.2 | 1.832 | 0.193 |
| Range | 70.0 – 125.0 | 70.0 – 130.0 |  |  |
| **Ankle brachial index** |  |  |  |  |
| Mean ± SD | 0.92 ± 0.16 | 1.03 ± 0.12 | 4.945 | < 0.001* |
| Range | 0.7 – 1.3 | 0.8 – 1.3 |  |  |

**Table 3.** Association between nephrotic syndrome and socio-demographic/ clinical characteristics.

| Variables | PAD (n = 44)<br>n (%) | No PAD (n = 56)<br>n (%) | $\chi^2$ | p-value |
| --- | --- | --- | --- | --- |
| **Age group (Years)** |  |  |  |  |
| 1 - 5 | 9(34.6) | 17(65.4) | 2.002 | 0.368 |
| 6 - 10 | 19(43.2) | 25(56.8) |  |  |
| 11 – 15 | 16(53.3) | 14(46.7) |  |  |
| **Gender** |  |  |  |  |
| Male | 24(41.4) | 34(58.6) | 0.385 | 0.535 |
| Female | 20(47.6) | 22(52.4) |  |  |
| **Social class** |  |  |  |  |
| Lower | 18(43.9) | 23(56.1) | 1.005 | 0.605 |
| Middle | 14(38.9) | 22(61.1) |  |  |
| Upper | 12(52.2) | 11(47.8) |  |  |
| Waist circumference | 64.89±10.4 | 59.16±7.2 | 3.268 | **0.001\*** |
| Hip circumference | 70.89±12.9 | 65.10±8.9 | 2.653 | **0.009\*** |
| Waist hip ratio | 0.92±0.1 | 0.91±0.1 | 1.115 | 0.269 |
| WAZ | 0.59±1.2 | 0.24±1.3 | 0.975 | 0.334 |
| HAZ | -1.13±1.2 | -1.01±1.3 | -0.383 | 0.703 |
| BAZ | 1.57±1.2 | 1.06±1.4 | 1.568 | 0.121 |
| RBG (mg/dl) | 86.18±20.5 | 85.34±14.1 | -0.243 | 0.809 |
| Serum protein(mg/dl) | 5.17±0.8 | 5.40±1.2 | -1.122 | 0.264 |
| Albumin (mg/dl) | 3.53±0.9 | 3.39±0.9 | 0.785 | 0.435 |
| Triglyceride (mg/dl) | 113.3 (82-157) | 105.1 (59-132) | -1.934 | **0.043\*** |
| VLDL (mg/dl) | 22.7 (16-31) | 21.0 (12-26) | -1.674 | **0.047\*** |
| TC (mg/dl) | 171.5 (146-269) | 156.7 (117-206) | -1.559 | **0.033\*** |
| LDL (mg/dl) | 110.3 (80-174) | 80.8 (52-138) | -2.024 | **0.043\*** |
| Duration since diagnosis | 32.0 (21.5-58.5) | 22.00 (11.0-48.0) | -2.286 | **0.022\*** |
| Cumulative steroid dose (mg) | 4605.0 (3775-7415) | 3675.0 (2925-4750) | -3.320 | **0.001\*** |
| Duration of steroid use (months) | 24.00 (11.0-48.0) | 12.00 (7.0-24.0) | -2.095 | **0.036\*** |
| Response to steroid |  |  |  |  |
| Steroid dependent | 2(66.7) | 1(33.3) | 17.537 | **<0.001\*** |
| Steroid resistance | 15(88.2) | 2(11.8) |  |  |
| Steroid sensitive | 27(33.8) | 53(56.0) |  |  |

**Table 4:** Independent predictors of peripheral arterial disease among patients with nephrotic syndrome.

| Variables | AOR | 95% C I | p-value |
| --- | --- | --- | --- |
| Age | 0.973 | 0.943-1.004 | 0.083 |
| Gender: Female | 1.742 | 0.706-4.303 | 0.229 |
| Social class: Lower | 1.024 | 0.416-2.523 | 0.958 |
| Duration since diagnosis (months) | 4.372 | 2.934-12.301 | **0.021\*** |
| Response to steroid: Steroid resistance | 12.546 | 3.280-47.992 | **<0.001\*** |
| Cumulative steroid dose (mg) | 1.434 | 1.083-1.902 | **0.007\*** |
| Waist circumference (cm) | 1.225 | 0.951 - 1.577 | 0.116 |
| Hip circumference (cm) | 0.867 | 0.692 - 1.086 | 0.214 |
| Triglyceride (mg/dl) | 1.007 | 0.992 - 1.021 | 0.366 |
| VLDL (mg/dl) | 1.043 | 0.893 - 1.304 | 0.481 |
| TC (mg/dl) | 0.985 | 0.956 - 1.016 | 0.340 |
| LDL (mg/dl) | 1.015 | 0.985 - 1.045 | 0.328 |

The mean systolic ankle pressure was significantly lower in cases than the comparative group (p < 0.001) whereas mean systolic brachial pressure was comparable between both groups (p = 0.193). ABI was significantly lower in nephrotic syndrome patients than the comparative group (p < 0.001). Forty-four of hundred NS patients had peripheral arterial disease (ABI range 0.7 – 1.3) giving a prevalence rate of 44%. – fig 1. The corresponding figures for the comparative group were six of hundred subjects with a prevalence rate of 6.0%. Thus the prevalence of PAD was significantly higher in participants with NS (p < 0.001). There is about 12 fold (95% CI = 3.712 - 35.312) odds of developing PAD in participants with NS as compared to the comparative group.

**Fig 1.**
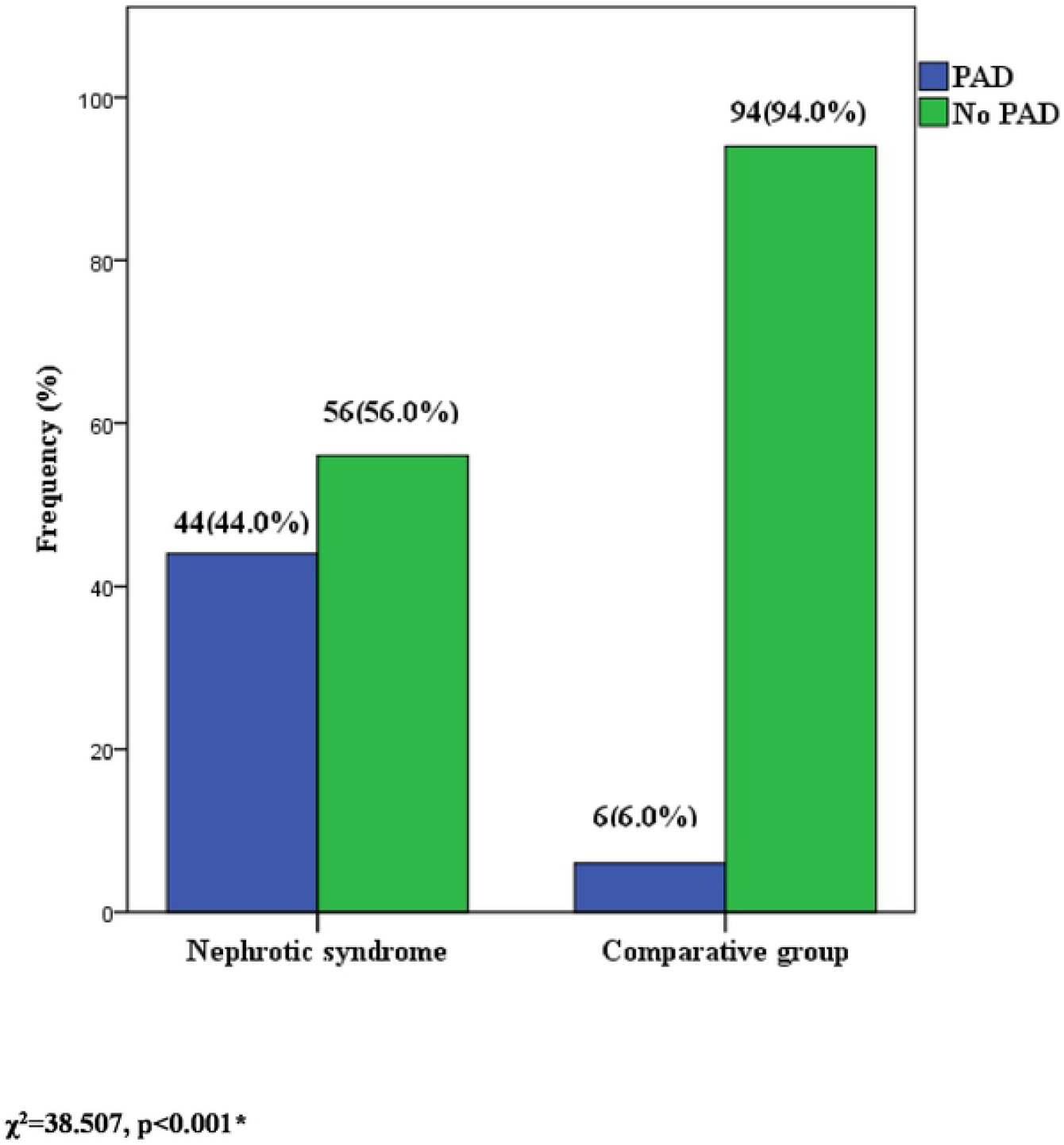
Prevalence of PDA in children with nephrotic syndrome compared with in apparently healthy comparative group.

The proportion of children with PAD was highest among the age group of 11-15 years. The occurrence of PAD did not differ significantly across gender and socio-economic status in participants with NS. Participants with PAD have significant higher waist circumference, hip circumference and lipid profiles than those without PAD. Duration since diagnosis, cummulative steroid dose and duration of steroid use were higher among participants with PAD (p < 0.05).

Table XI shows the result of a multiple logistic regression model that was developed to identify independent predictors of peripheral arterial disease. Increasing duration since diagonosis of NS (AOR=4.372; 95% CI= 2.934-12.301), steroid resistance (AOR=12.546; 95% CI = 3.280-47.992) and increasing cummlative steroid dose (AOR = 1.434; 95% CI = 1.083-1.902) are indepndent predictor of PAD.

## DISCUSSION

The present study among children with NS demonstrates a high prevalence of PAD, one of the least studied but potentially fatal complication of NS. Children with NS have about twelve fold odds of developing PAD than children without NS. This pattern was more prominent in those with steroid resistant NS than those responsive to steroid. High rate of PAD in children with steroid resistant NS has been previously documented by Mohammed et al.[4] The higher risk of PAD in steroid resistant NS may be attributed to persistence of proteinuria and progression to chronic kidney disease which is an established risk factor for PAD.[8,38] Chronic kidney disease leads to development of PAD through persistent proteinuria, endothelial dysfunction and chronic inflammatory state thereby enhancing plaque formation in the lumen of vessels.[12]

In the current study occurrence of PAD was found to increase with age albeit not significant. The few earlier studies in children with NS did not check for association of age with PAD. The finding of PAD occurring more among the older age subjects with NS in the current study is consistent with earlier reports in adults.[10,16,39] Changes in the structure and functions of large and small arteries with resultant loss of compliance in the vasculature increases with age.[40] The loss of compliance of these vessels encourages endothelial damage and plaque accumulation with resultant peripheral arterial disease.[16]

The majority of subjects with NS recruited in the current study were from the low social class as compared to the middle and upper class (41%, 36% and 23% respectively). This is consistent with findings from other African countries and Europe.[4,15,41] However, the proportion of children with PAD in the same cohort of NS subjects recruited was higher in the upper socio-economic group in the present study. This contrasts findings in the literature from developed countries where PAD was more common among the low socio-economic class.[42] The reason for this finding may be attributed to the fact that children from the higher social class in our environment are more likely to live a sedentary lifestyle with a high tendency for development of obesity. Also dietary intake of junks from affluence may be contributory.

In the current study, a significant proportion of children with PAD have had NS for more than twelve months. This finding is similar to what was found in Egypt among children with NS.[4] The duration of nephrotic hyperlipidaemia appears to be critical to initiating vascular and endothelial damage which favours influx of lipoprotein into the mesangium and ultimately leading to proliferation and sclerosis of the vessels. Lipoproteins are more elevated in children with long standing nephrotic syndrome than those who are recently diagnosed and thus increasing their risk for the development of peripheral arterial disease.[4,43] Screening of children who have had NS more than twelve months for PAD is thus highly recommended.

In the current study, chronic use of steroids was significantly associated with PAD. This is in keeping with a study in France among similar population of subjects by Willenberg et al.[44] Steroids have been reported to increase arterial calcification and its association with distal occlusive pattern has been observed in patients with chronic kidney disease.[44] In addition, chronic use of steroids may result in the development of hypertension which is a risk factor for PAD.[45] Children with NS who have frequent relapses requiring chronic use of steroids and those who are dependent on steroids should have more frequent checks of their ankle brachial index to check for PAD.

The current study found a significant number of subjects with PAD had a higher mean value of waist and hip circumference compared to subjects without PAD. This is difficult to compare with studies in children with NS as association with PAD was not checked in these studies. However the finding is similar to that of Martha et al [46] and Umueri et al [47] in adult subjects. Waist and hip circumference provides a unique indicator of body fat distribution in children. [48] It has been proven to be a surrogate marker of abdominal fat mass because it correlates well with subcutaneous and intra-abdominal fat mass.[48] Waist and hip circumference can identify patients who are at increased risk of obesity-related cardio-metabolic disease including PAD above and beyond the measurement of body mass index.[48,49] Combined measurements of waist circumference, hip circumference and waist to hip ratio are reported to be better predictors of cardiovascular disease including PAD in children than body mass index.[50] Obesity has been linked closely with other risk factors for the development of PAD like hyperlipidaemia, hypertension and poor glycaemic control which could lead to endothelial dysfunction and ultimately PAD. Routine measurement of waist circumference, hip circumference and waist to hip ratio will be highly recommended for children with nephrotic syndrome during clinic visits to identify early deviation from normal.

The mean value of weight-for-age Z score was higher in the group with PAD when compared to those without PAD in the current study. This finding may likely be due to the effect of prolonged use of steroids which is notorious for inducing hypercortisolism related side effects. High cortisol levels can lead to increased appetite, truncal fat accumulation, and altered lipid and glucose metabolism leading to obesity.[50] Similarly, the mean value of height-for-age Z score was lower in the group with PAD than in the group without PAD. This may also be due to the effect of prolonged steroid use. A comparable finding was reported by Simmonds et al [51] among children with steroid dependent nephrotic syndrome. Steroids have inhibitory effects on growth hormone release. It also has direct effect on growth plate by suppressing chondrocyte proliferation, matrix proteoglycan synthesis and mineralization.[51] Moreover, persistence and or relapse of NS dictates continued use of steroids. Thus, simultaneously chronic disease state of NS may predispose to PAD and prolonged steroid use to impaired linear growth.

One of the findings of the present study is that the average values of serum lipids (total cholesterol, triglycerides, very low density lipoprotein and low density lipoprotein) were significantly higher in subjects with PAD than those without PAD. These results are in agreement with an Egyptian study also among children with NS.[4] Hyperlipidaemia increases the risk of premature atherosclerosis.[4] Infiltration and retention of low density lipoproteins in the arterial wall is a critical event that sparks an inflammatory response which results in the formation of plaques that leads to PAD.[52] The elevated average values of serum lipids in this current study is also consistent with findings in children and adults with nephrotic syndrome in Nigeria.[53,54] Hyperlipidaemia in NS results from a compensatory response to reduced serum protein that leads to increased synthesis and decreased catabolism of lipids.[33] It is advocated that serum lipid profile of children with nephrotic syndrome be done routinely and more frequently in those who have frequent relapses and are steroid resistant.

In the present study, the mean values of both serum protein and albumin were lower in the subjects with peripheral arterial disease. This is comparable to findings in children and adults.[4,55] With proteinuria and resultant hypoproteinaemia there is a compensatory hyperlipidaemia which can lead to endothelial damage and increase the risk of developing peripheral arterial disease.

In the current study, duration since diagnosis of nephrotic syndrome, steroid resistant nephrotic syndrome and cumulative dose of prednisolone greater than 3000mg were found to be independent predictors of peripheral arterial disease. Close monitoring of children with nephrotic syndrome with long duration since diagnosis and those with steroid resistant NS is important before they develop PAD. Prompt referral of children with NS who have abnormal ABI to the cardiologist for follow up is also advocated especially before they develop life threatening complications.

## Data Availability

All information and files will be available after acceptance for publication. XXXXXXXX

## Acknowledgments

The authors acknowledge Dr Ayo Faremi for his help with procuring some of the devices and accessories used for the study. Our thanks also goes to Drs Mariam Disu, Amontunur Lamina, Oluwabukola Kuti, Ibukunoluwa Adeboye and Tracy Ossai for their help with sample and data collection. The amiable and enthustiatic subjects along with their parents/guardians who participated in the study are specially appreciated.

